# Governing Decisions of Probability Cutoffs in Clinical AI Deployment: A Case Study of Asthma Exacerbation Prediction

**DOI:** 10.64898/2026.03.18.26348562

**Authors:** Lu Zheng, Bhavani Singh Agnikula Kshatriya, Joshua W. Ohde, Lauren Rost, Momin M. Malik, Kevin J. Peterson, Tracey Brereton, Brenna T. Loufek, Tara V. Pereira, Chenyu Gai, Miguel A. Park, Martha F. Hartz, Joy Fladager-Muth, Chung Wi, Cui Tao, Vesna Garovic, Young Juhn, Shauna M. Overgaard

## Abstract

Models that estimate the probability of an adverse clinical outcome require an operational cutoff to translate continuous estimated probabilities into discrete labels that can trigger clinical action. Although statistical methods identify ‘optimal’ cut-offs, threshold selection ultimately reflects value judgments regarding harm tolerance, resource allocation, and workflow feasibility. We describe a governance-informed approach to selecting a deployment threshold for an asthma exacerbation (AE) prediction model integrated into clinical workflows. Using prevalence-adjusted performance metrics and real-world provider capacity modeling, we evaluated multiple candidate thresholds and quantified downstream workload and missed-event trade-offs. We demonstrate that statistically optimal thresholds may produce operationally infeasible alert volumes or unacceptable miss rates. We propose a structured threshold governance framework integrating statistical performance, clinical utility, stakeholder input, and human oversight safeguards. This case illustrates how threshold decisions should be treated as organizational governance processes rather than purely technical optimizations.

## Introduction

Clinical artificial intelligence (AI) models are increasingly embedded within electronic health record systems to support proactive patient care, and 65% of U.S. hospitals used predictive AI models.^1^ While model development studies emphasize discrimination and calibration metrics, real-world deployment requires converting continuous probability estimates into actionable clinical interventions. This conversion is necessary because probability scores alone do not directly inform clinical decisions. A threshold must be determined to inform when a predicted risk is high enough to justify downstream intervention. Setting this threshold translates the model’s output into a binary clinical decision such as whether to alert providers, initiate additional monitoring, or escalate care, which ensures that predictive analytics are meaningfully integrated into clinical workflows. The choice of threshold reflects a balance between maximizing patient benefit, minimizing unnecessary interventions, and aligning with operational resource constraints.^2^

Despite its importance on clinical outcome, threshold selection is often treated as a technical consideration that relies on receiver operating characteristic (ROC) curve optimization or F1 score maximization. However, it is important to note that routinely selecting thresholds based on ROC curve metrics or overall accuracy assumes equal error costs and consistent prevalence, which are often not valid in real clinical settings.^3^ Although decision curve analysis is a well-regarded method for quantifying net benefit, it should be used to assess clinical utility across various thresholds rather than to identify a single ‘optimal’ cutoff.^4^ In truth, there is no universally optimal threshold, as statistical standards do not consider practical feasibility, provider workload, acceptable risks, or organizational priorities. Despite the importance of cutoff choice at deployment, explicit guidance on how health systems should select, justify, and document operational cutoffs remains limited and fragmented. Existing literature more often emphasizes technical performance optimization or decision-analytic evaluation than the institutional governance processes by which these cutoffs are chosen in real clinical settings.

Integrating predictive models into clinical workflows requires evaluating their financial and physical impact on patients, minimizing harm and resource use. Institutions should account for workload from false positives and negatives, as these affect staff capacity. The required resources and patient outcomes depend on the intervention triggered by the model, while clinicians’ preferences and behaviors shape response to model outputs. Threshold selection should consider all these factors to enhance adoption and improve care. Consequently, threshold determination is frequently conducted in an informal and fragmented manner, with limited documentation by both developers and clinicians. In contrast to this, regulatory authorities, including the FDA, NIST, and the Coalition for Health AI, consistently emphasize the importance of documenting model performance and providing clinical justification at specified thresholds. To address this gap, governance processes must ensure that policy is translated into structured documentation, explicitly detailing threshold rationale, performance metrics, and workflow implications. This paper presents a real-world case study of threshold selection for an asthma exacerbation prediction model in pediatric care, aiming to encourage the development of generalizable structured documentation that supports transparent decision-making and streamlines governance review.^5,6^

## Methods

To operationalize transparent threshold selection for a predictive model, we implemented a structured deliberation process supported by a standardized documentation template. The approach was designed to align with emerging governance expectations that clinical AI systems document the rationale, performance characteristics, and workflow implications associated with operational decision thresholds.

The process was applied to an asthma exacerbation (AE) risk prediction model integrated into a clinical decision support interface. The model generates a continuous risk score indicating the probability that a patient will experience an asthma exacerbation within the next 12 months. Patients with scores exceeding a selected threshold are flagged as high risk, prompting follow-up review and outreach by care teams through the clinical workflow. We implemented a three-step framework for operational threshold selection consisting of candidate threshold identification, structured clinical deliberation, and governance documentation (Figure 1).

**Figure 1.**
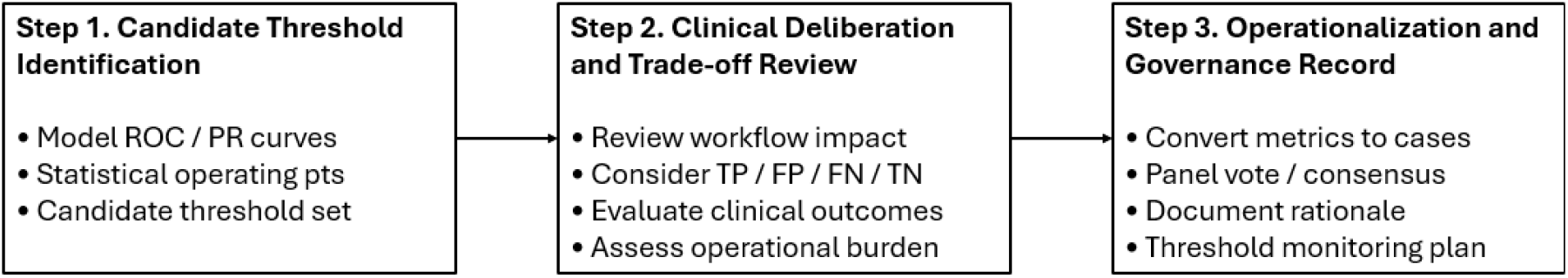
Framework for governance-informed threshold selectionThree-step process for selecting an operational model threshold: identify candidate cutoffs, evaluate clinical and workflow implications, and document the final decision for governance and monitoring.

A facilitated deliberation session was conducted with four practicing clinicians serving as an advisory panel. The objective of the session was to evaluate candidate operating thresholds and select a threshold appropriate for clinical deployment. Prior to discussion, the facilitator presented a structured set of materials designed to support informed decision making. First, the clinical workflow triggered by the model output was introduced to clarify how flagged patients would be reviewed and managed in practice. Next, the facilitator described the clinical and operational implications of the four classification outcomes (true positives, false positives, true negatives, and false negatives) and asked panelists to qualitatively consider the relative clinical consequences and resource implications associated with each outcome category.

The facilitator then presented several candidate thresholds along the model’s receiver operating characteristic (ROC) curve. Candidate cuttofs included thethe statistical maximum F1-score point, the Youden’s index point (defined as the cutoff maximizing sensitivity + specificity – 1) and nearby ROC operating points selected to illustrate clinically relevant trade-offs among sensitivity, positive predictive value, and flag rate.

To contextualize these metrics within institutional practice, model performance metrics were recalculated using the historical asthma exacerbation prevalence of 24.1% observed in the feasibility cohort. Model rates were translated into projected annual case counts using local operational data consisting of 1,235 asthma patients managed by 167 providers over a one-year period. Estimated numbers of patients captured, missed exacerbations, and total flagged cases were calculated for each threshold scenario.

Following review of these scenarios, panelists discussed the trade-offs between identifying patients at risk and maintaining an operationally feasible workload. Panelists then voted on their preferred threshold option, and consensus was reached through facilitated discussion. The rationale, evaluated thresholds, and expected operational impact were documented using a structured threshold documentation template to support governance review and future monitoring.

## Results

### Thresholds Performance Evaluation

Five candidate operating thresholds were evaluated, including the statistical maximum F1-score point and four neighboring thresholds along the receiver operating characteristic (ROC) curve. The additional candidate cutoffs were selected to span clinically meaningful trade-offs in sensitivity, positive predictive value, and flag rate (Figure 2). Sensitivity across thresholds ranged from 58.1% to 97.1%, while positive predictive value (PPV) ranged from 44.3% to 26.1%. Corresponding flag rates varied substantially from 31.6% to 89.2% of the patient population, illustrating the operational consequences of threshold selection.

**Figure 2.**
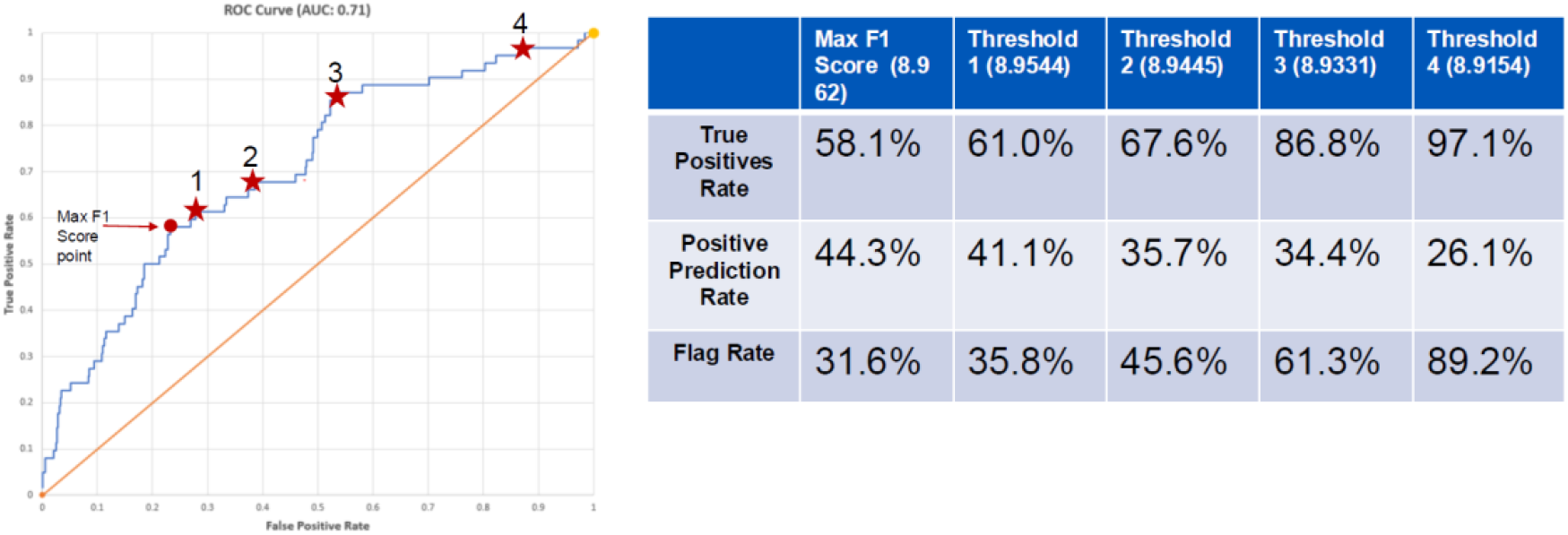
ROC-based candidate thresholds for asthma exacerbation risk prediction. ROC curve with five candidate thresholds considered for deployment, including the maximum F1-score point, the Youden index point, and nearby alternatives showing key performance trade-offs.

To support clinical interpretation, model performance was contextualized using real-world operational data from the feasibility cohort. The cohort included 1,235 pediatric asthma patients managed by 167 providers over a one-year period, with an observed asthma exacerbation prevalence of 24.1% (298 events). It was confirmed by the panelist that approximately a pediatrician would see two asthma patients each month on average.

Translating statistical metrics into operational case counts revealed substantial differences in downstream workload and missed-event rates. At the most conservative operating point near the maximum F1-score threshold, the model would correctly identify 173 of 298 exacerbation events, while 390 patients (31.6%) would be flagged for clinical review. In contrast, the lowest threshold scenario captured 289 of 298 exacerbations but would require follow-up review for 1,103 patients (89.2%). Intermediate thresholds produced graduated trade-offs between these extremes. For example, one candidate threshold captured 182 exacerbations while generating 442 flagged cases (35.8%), whereas another captured 258 exacerbations but required review of 756 patients (61.3%).

These comparisons demonstrated that small adjustments in the risk score threshold produced large differences in both clinical detection and operational burden. In particular, thresholds that maximized statistical sensitivity substantially increased the number of patients requiring follow-up evaluation, potentially exceeding feasible provider capacity.

### Deliberation Outcomes

During the facilitated discussion session, clinicians reviewed the operational projections associated with each candidate threshold and evaluated them in relation to the anticipated clinical workflow triggered by the model. Panelists emphasized that the intervention associated with the asthma exacerbation prediction model primarily involved non-invasive follow-up activities, such as chart review and patient outreach, while asthma exacerbation is a life-threatening event. As a result, clinicians expressed more tolerance for false positives (right-shifting threshold) but remained concerned about excessive alert volume that could lead to alert fatigue and reduced trust in the model.

Discussion centered on balancing two competing priorities: maximizing the identification of patients at risk for exacerbation and maintaining a manageable workload for care teams responsible for follow-up. Panelists noted that extremely sensitive thresholds would generate alert volumes that could overwhelm clinical staff and dilute attention to the highest-risk patients. Conversely, overly restrictive thresholds would fail to identify a meaningful proportion of preventable exacerbation events.

Through facilitated discussion and voting, the panel reached consensus on threshold 3 that balanced event detection with operational feasibility. This operating point captured a majority of exacerbation events while limiting the proportion of flagged patients to a level considered manageable for individual pediatricians within existing care team workflows.

### Governance Documentation of the Threshold Decision

The threshold decision and supporting rationale were subsequently documented using the structured governance template proposed in this study (Table 1). The template illustrates how the asthma exacerbation threshold selection process can be translated into a standardized governance record.

**Table 1.**
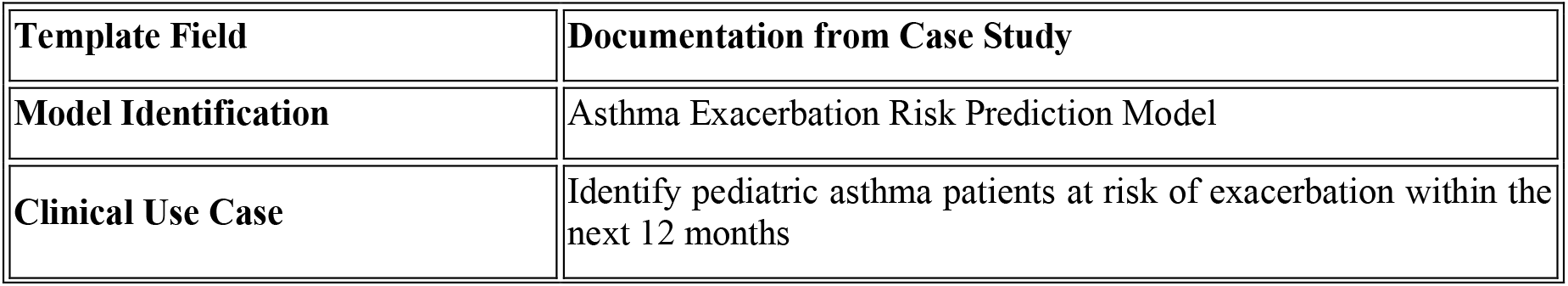
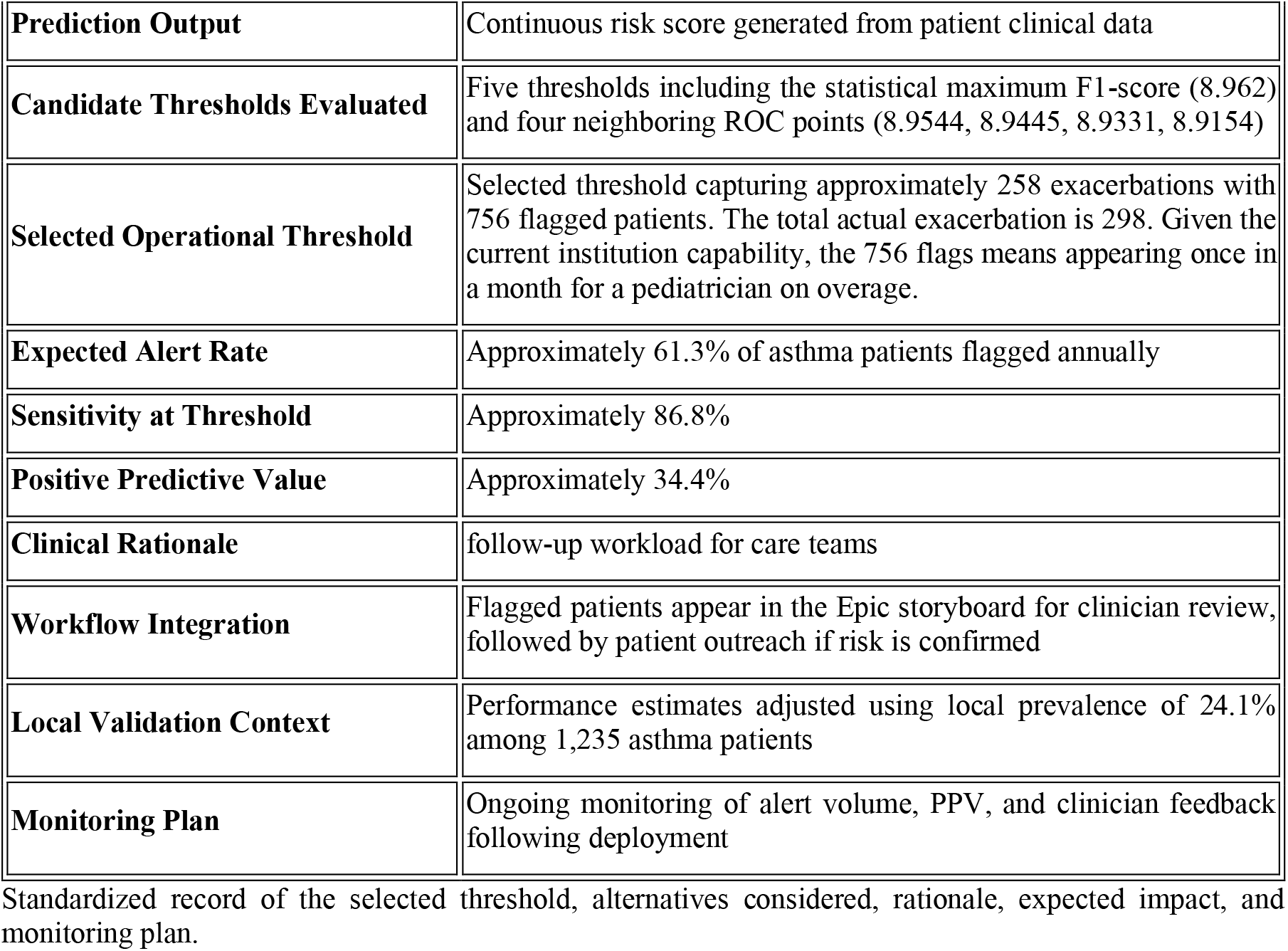
Governance documentation for threshold selection.

## Discussion

This case study highlights that the selection of operational thresholds for clinical AI models cannot be treated as a purely statistical optimization problem. Although model development commonly reports discrimination metrics such as AUROC and identifies “optimal” cutoffs using statistical criteria such as maximum F1-score or ROC proximity to the top-left corner, these operating points may not align with real-world clinical and operational constraints. In our analysis, small changes in the threshold value produced substantial differences in both the number of exacerbation events captured and the volume of patients flagged for follow-up, as shown in Figure 2. These differences translate directly into variations in provider workload and potential alert fatigue, demonstrating that threshold decisions have meaningful implications for clinical workflow and resource utilization.

The structured deliberation process revealed that clinicians evaluate model outputs through the lens of downstream intervention burden and clinical risk. In this case, the primary intervention triggered by a flagged prediction involved relatively low-risk activities such as chart review and follow-up phone outreach. As a result, clinicians expressed greater tolerance for false positive alerts because the associated costs were limited to additional provider effort and minor inconvenience to patients. In contrast, false negative predictions were perceived as significantly more harmful because missed exacerbation risks could result in emergency department visits, hospitalizations, or deterioration in patient health. This asymmetric weighting of prediction errors influenced the panel’s preference for a threshold that prioritized detection of exacerbation events while maintaining a manageable level of operational workload.

Importantly, the facilitated discussion demonstrated that translating model performance metrics into projected patient counts substantially improved stakeholders’ ability to evaluate threshold trade-offs. Discussion centered on balancing two competing priorities - maximizing identification of patients at risk for exacerbation and maintaining a manageable workload for care teams responsible for follow-up (Figure 3).

**Figure 3.**
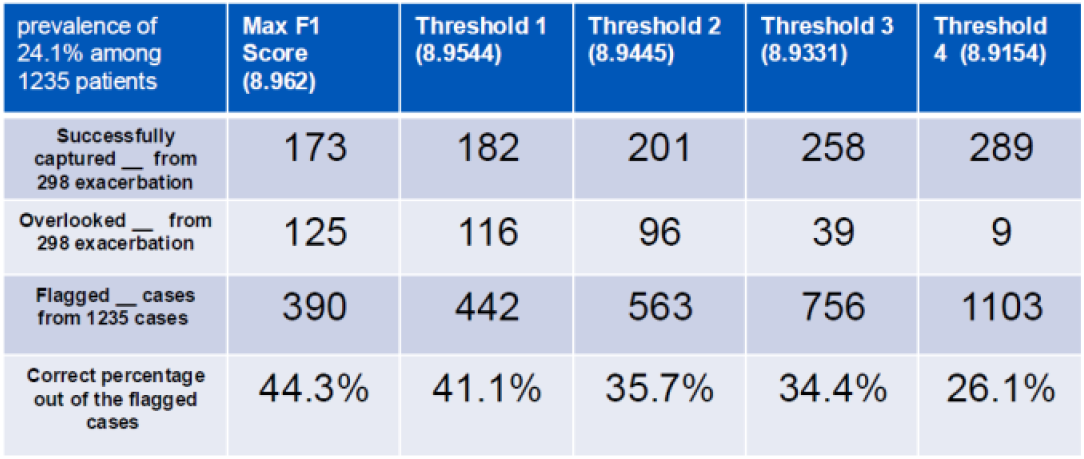
Projected clinical consequences of candidate deployment thresholds. ROC-based candidate thresholds considered for deployment, with corresponding projections of exacerbations identified, missed events, and patients flagged to show the operational impact of threshold choice.

Presenting sensitivity and positive predictive value alone provided limited intuition for clinical decision makers. However, converting these metrics into estimated numbers of captured exacerbations, missed events, and flagged patients grounded the discussion in real-world operational consequences. This approach enabled clinicians to assess whether a proposed operating point would be feasible within existing care team capacity.

Beyond the specific case presented here, the study highlights a broader governance gap in clinical AI implementation. While regulatory guidance and emerging AI governance frameworks emphasize the importance of documenting model performance at the chosen operating threshold, the decision-making process used to select that threshold is often informal and poorly documented. This relative lack of explicit guidance for selecting and documenting deployment cutoffs may help explain why these decisions are often made informally, even though they materially affect workload, missed events, and patient safety. Without explicit documentation of the rationale and operational assumptions underlying threshold choices, governance committees may lack the information necessary to evaluate deployment decisions or monitor system performance after implementation. The structured documentation template introduced in this study addresses this challenge by translating threshold selection into a reproducible governance artifact (Table 1). By capturing candidate thresholds, performance metrics, clinical rationale, workflow implications, and monitoring plans, the template provides a concise summary that can support governance review and facilitate future model updates. This documentation also promotes transparency by making explicit the value judgments and trade-offs embedded in threshold decisions.

Several limitations should be considered when interpreting the findings. First, the clinician deliberation involved a relatively small advisory panel and may not fully represent the perspectives of all stakeholders involved in asthma care. Second, operational projections were derived from historical prevalence and workload assumptions, which may change over time as clinical practices evolve or as model performance shifts following deployment. Finally, the case study focuses on a single predictive model within one institutional context. Therefore, the specific threshold selected should not be interpreted as universally applicable. Nevertheless, the governance approach demonstrated here is generalizable and may be adapted to other predictive models and clinical workflows.

Future work could expand this framework by incorporating formal decision-support methods, such as decision curve analysis or cost–benefit modeling, into structured deliberation sessions. In addition, integrating threshold documentation directly into AI governance intake workflows may further streamline review processes and ensure consistent evaluation across multiple AI deployments.

## Conclusion

Operational threshold selection is a central yet often under-documented decision in the deployment of clinical AI systems. While statistical metrics can identify candidate operating points, the final threshold must ultimately reflect clinical priorities, workflow feasibility, and organizational risk tolerance. This case study demonstrates how structured clinical deliberation, combined with operational modeling and standardized documentation, can support transparent and accountable threshold decisions. By treating threshold selection as a governance process rather than a purely technical optimization task, health systems can better align predictive model deployment with patient safety, clinical workflow realities, and responsible AI oversight.

## Data Availability

All data produced in the present study are available upon reasonable request to the authors.

